# Evidence for feasibility of mobile health and social media-based interventions for early psychosis and clinical high risk

**DOI:** 10.1101/2022.04.01.22273303

**Authors:** Olivia H. Franco, Monica E. Calkins, Salvatore Giorgi, Lyle H. Ungar, Raquel E. Gur, Christian G. Kohler, Sunny X. Tang

## Abstract

**Background:** Digital technology, the internet and social media are increasingly investigated as a promising means for monitoring symptoms and delivering mental health treatment. These apps and interventions have demonstrated preliminary acceptability and feasibility, but previous reports suggests that access to technology may still be limited among individuals with psychotic disorders relative to the general population.

**Objective:** We evaluated and compared access and use of technology and social media in young adults with psychotic disorders (PD), clinical risk for psychosis (CR), and psychosis-free youths (PF).

**Methods:** Participants were recruited through a coordinated specialty care clinic dedicated towards early psychosis as well as ongoing studies. We surveyed 21 PD, 23 CR, and 15 PF participants regarding access to technology and use of social media, specifically Facebook and Twitter. Statistical analyses were conducted in R. Categorical variables were compared among groups Fisher’s exact test, continuous variables were compared using one-way ANOVA, and multiple linear regressions were used to evaluate for covariates.

**Results:** Access to technology and social media were similar among PD, CR and PF. Individuals with PD, but not CR, were less likely to post at a weekly or higher frequency compared to psychosis-free individuals. We found that decreased active social media posting was unique to psychotic disorders and did not occur with other psychiatric diagnoses or demographic variables. Additionally, variation in age, sex, Caucasian vs. non-Caucasian race did not affect posting frequency.

**Conclusions:** For young people with psychosis spectrum disorders, there appears to be no “technology gap” limiting the implementation of digital and mobile health interventions. Active posting to social media was reduced for individuals with psychosis, which may be related to negative symptoms or impairment in social functioning.

## Introduction

The increasing availability of technology has opened a new avenue for making healthcare more accessible to a broader population and for the development and implementation of digital interventions. In recent years, mobile and web-based health interventions have been used with great promise to assess and coordinate with mental health clients [1]. Social media and mobile health interventions present a unique opportunity to engage young people experiencing the emergence of a psychotic disorder (PD) or at clinical risk (CR) for psychosis - when there are clinically significant symptoms of psychosis, but threshold criteria are not yet met. The importance of early intervention and prevention in psychosis is a key factor in long term success, and it has been shown that decreasing the duration of untreated psychosis increases long-term treatment outcome [2,3].

Mobile and social media interventions and assessment tools are relatively new in psychiatry, but initial studies show promising engagement, efficacy, and prognostic utility [4,5]. For example, a social media language-based classification model was able to identify early relapse signs for people experiencing first episode psychosis [6]. Relapse events were also predicted in another study by changes in smartphone-enabled social activities [7]. Among interventions, patients with PD benefitted from a combined treatment protocol involving monitoring via mobile health interventions and psychosocial services. Digital social networks have been integrated into mobile health apps to facilitate peer-support and improve amotivation symptoms [8]. Other promising mobile health interventions include reminders for medication adherence and appointments, interventions, and case management via text messages and applications on a smartphone. Mobile interventions can also vary from standardized communication to live interactions with clinicians; for example, the patient can have live sessions with a therapist via video conferencing or text messaging, or they can receive standardized communications like pre-recorded videos, and tip sheets based on their input at the time of communication [9].

Access to technology among the target population is critical for the feasibility of mobile and social media-based interventions. Globally, access to technology has increased dramatically across all age groups, but a “technology gap” may exist for individuals at risk and suffering from psychotic disorders [2]. Among adult patients recruited from an inpatient and outpatient clinic, only 48% have access to the internet, and only 27% used social networking sites on a daily basis [10]. However, younger age may be associated with greater access to technology. In one study of young people in their first episode of psychosis, 100% used social media, and 90% of those reported daily usage for an average of about 2 hours daily [5]. Another study completed in Spain showed that first-episode patients had similar interest in but decreased access to digital technologies including computers and smartphones [11]. If young people struggling with psychosis commonly access digital technology and social media, it would provide an avenue not only for outreach, but also a method of monitoring symptoms and side effects and providing treatment.

To our knowledge, no study has directly compared access and use of technology and social media across the psychosis spectrum among CR, PD, and PF youths. Most studies also do not distinguish between passive access of social media and active posting. In this study, we surveyed young adults with psychosis, at clinical risk for psychosis, and psychosis-free comparison participants about their access to digital technology, the internet, and both access and posting to Facebook and Twitter. We hypothesized that the “technology gap” is decreased or absent among young adults with psychosis and clinical risk, suggesting that social media and mobile health interventions would be feasible and implementable in this population.

## Methods

Three groups were evaluated: young adults with psychotic disorders (PD), clinical risk for psychosis (CR), and psychosis-free youths (PF). A total of 59 participants aged 18-32 years were included (Table 1). PD participants were recruited from the University of Pennsylvania Psychosis Evaluation and Recovery Center (PERC), a coordinated specialty care clinic dedicated towards early psychosis. CR and PF participants were recruited among participants of other ongoing studies at the University of Pennsylvania and Children’s Hospital of Philadelphia Lifetime Brain Institute (LiBI). We were interested in the main effect of psychosis on access to technology and use of social media. Because mental health disorders are common in the community for young adults and adolescents [12], our psychosis-free comparison group did not exclude people with non-psychotic disorders.

**Table 1.** Sample Characteristics, Access to Technology and Social Media Use.

| Variable | PF | CR | PD | p |
| --- | --- | --- | --- | --- |
| **n** | 15 | 23 | 21 |  |
| **Age (mean ± SD, yrs.)** | 23.5 ± 4.2 | 22.0 ± 2.9 | 24.2 ± 3.4 | 0.10 |
| **Sex** |  |  |  | 0.01 |
| **Female (n, %)** | 11 (73%) | 12 (52%) | 5 (24%) |  |
| **Male (n, %)** | 4 (27%) | 11 (48%) | 16 (76%) |  |
| **Race** |  |  |  | 0.09 |
| **Caucasian (n, %)** | 4 (29%) | 11 (50%) | 14 (67%) |  |
| **African American (n, %)** | 7 (50%) | 9 (41%) | 6 (29%) |  |
| **Mixed (n, %)** | 1 (7%) | 2 (9%) | 0 (0%) |  |
| **Asian (n, %)** | 2 (14%) | 0 (0%) | 1 (4%) |  |
| **Other (n, %)** | 0 (0%) | 0 (0%) | 0 (0%) |  |
| **Comorbidities** |  |  |  |  |
| **ADHD (n, %)** | 2 (13%) | 3 (13%) | 2 (10%) | 1.00 |
| **Anxiety Disorder (n, %)** | 2 (13%) | 15 (65%) | 5 (24%) | 0.002 |
| **Mood Disorder (n, %)** | 3 (20%) | 12 (52%) | 7 (33%) | 0.14 |
| **Access to Technology** |  |  |  |  |
| **Mobile phone** | 15 (100%) | 22 (96%) | 21 (100%) | 1.00 |
| **Smartphone** | 15 (100%) | 22 (96%) | 20 (95%) | 1.00 |
| **Computer** | 13 (87%) | 21 (91%) | 20 (95%) | 0.84 |
| **Internet** | 15 (100%) | 23 (100%) | 20 (95%) | 0.61 |
| **Social Media Use** |  |  |  |  |
| **≥ Weekly access** | 10 (67%) | 17 (74%) | 13 (62%) | 0.73 |
| **Facebook** | 10 (67%) | 15 (65%) | 13 (62%) |  |
| **Twitter** | 3 (20%) | 5 (22%) | 0 (0%) |  |
| **≥ Weekly posting** | 4 (27%) | 10 (43%) | 1 (5%) | 0.01 |
| **Facebook** | 4 (27%) | 9 (39%) | 1 (5%) |  |
| **Twitter** | 2 (13%) | 3 (13%) | 0 (0%) |  |
| **Facebook (ever)** | 14 (93%) | 17 (74%) | 15 (71%) | 0.27 |
| **Twitter (ever)** | 4 (27%) | 6 (26%) | 2 (10%) | 0.30 |

Note: PF – Psychosis-Free; CR – Clinical risk; PD – Psychotic disorder.

All stable outpatients at PERC aged 18-35 years as well as potential participants in LiBI studies were approached and asked for their consent to participate in this survey as a screening procedure for larger studies. Participants all resided in the greater Philadelphia area which includes both an urban and the surrounding suburban environment. There was no remuneration for completing this survey. Several eligible individuals were not interested in research in general, but no one specifically refused to participate in this survey. This was a brief single timepoint assessment and no one withdrew prior to its completion. All procedures were approved by the IRB at the University of Pennsylvania.

The survey was completed verbally with a research coordinator either in-person or over the telephone. Individuals who agreed were surveyed via a questionnaire regarding their access to technology and use of social media (Facebook and Twitter). Access to technology was defined generally as the ability to use the technology in a dependable manner, including personal ownership, shared devices, and public access points. Active posting was defined as commenting, posting status updates, or otherwise contributing original content, versus passive access of social media, which includes scrolling, viewing, or liking. We did not distinguish between accessing the internet via Wifi or a data plan.

The PF and CR groups underwent semi-structured interviewing with the Structured Interview for Prodromal Syndromes (SIPS), which assessed threshold and subthreshold symptoms of psychosis. Determination for CR or PF status was made by consensus case conference. The PD group underwent consensus clinical diagnosis by psychiatrists and psychologists specializing in psychotic disorders and were determined to have one of the following DSM 5 psychotic disorders: schizophrenia (n=13), schizoaffective disorder (n=4), bipolar I disorder with psychotic features (n=1), or unspecified psychotic disorder (n=3). Co-morbid nonpsychotic disorders were evaluated based on DSM 5 criteria and were also present in all three groups, in proportion to expected population rates (Table 1).

Statistical analyses were conducted in R, v. 3.5.2. Categorical variables were compared among groups using Fisher’s exact test, include the main outcomes of technology access and posting frequency versus group. Fisher’s exact test was also used to compare social media access and posting rates for other diagnoses as well as for sex and race effects. Age differences among the groups and between active vs. non-active users was compared using one-way ANOVA. To account for group differences in age, sex, and race, we additionally performed logistic regressions predicting social media access and posting with group as the independent variable of interest, and covarying for the demographic variables. Significance was two-tailed with α=0.05.

## Results

Table 1 displays details for 21 PD, 23 CR, and 15 PF participants. There was uniformly high access to mobile phones (96%-100%, p=1.00), smartphones (95%-100%, p=1.00), computers (85%-95%, p=0.84), and the internet (95%-100%, p=0.61). The majority of young adults accessed Facebook, but not Twitter. Social media access rates were similar for all three groups (62%-74% at least weekly, p=0.73). However, there was a significant main group effect for social medial posting (5%-42% at least weekly, p<0.01). CR actively posted at a similar rate compared to psychosis-free individuals (CR 43% vs. PF 27%, p=0.31), but PD actively posted at a significantly lower rate than the non-psychotic groups (5%, p<0.01). When examining Facebook usage alone, as this was the more population platform, we found consistent results: access was not affected by psychosis group (p=1.00), but there was a significant main of effect for group on posting (p= 0.02). While covarying for sex, age, and race, psychosis diagnosis remained a significant predictor of decreased active social media posting (Std. β=-1.22, p=0.03) but not a significant predictor for social media access (Std. β=-0.29, p=0.40).

Decreased active social media posting was unique to psychotic disorders and there was no group effect for diagnosis with ADHD (p=0.67), mood (p=0.54), or anxiety disorders (p=1.00) when comparing prevalence of weekly or greater use. Variation in age (p=0.63), sex (p=0.13), and Caucasian vs. non-Caucasian race (p=0.77) did not affect posting frequency.

## Discussion

We found that youths with PD and CR who participate in our clinical care and research programs have similar access to technology and use social media to a similar degree compared with PF individuals. This supports the feasibility of mobile health and social media interventions in young CR and PD populations in terms of access to technology. Our results were consistent with those found in previous studies which showed that the use of digital technology in the treatment of people with psychosis spectrum disorder and other chronic serious mental illnesses is a viable method of delivery of services. In our study, the absolute rate of mobile phone and smartphone ownership and internet usage was higher than those reported by Young et al., likely because of the younger age of our participants [13]. Most previous studies have compared access to technology between mental health clients and published normative data for the general population but have not directly compared across groups using a consistent standardized methodology. One study directly compared the use of (not access to) technology between first-episode psychosis patients and healthy control subjects; they found significant but small-moderate effects of decreased frequency of use for computers, tablets, smartphones, and Smart TV (but not game consoles) [11]. However, it is unclear whether demographic differences may have accounted for some of the disparity. No studies have included a psychosis clinical risk comparison group.

Another relevant consideration is comfort with technology among clinicians. A recent study by Camacho and Torous surveyed 42 Coordinated Specialty Care (CSC) clinics and found that health care providers were supportive of implementing technology in their care model for early psychosis; while 69% of surveyed staff was confident in their ability to provide technical assistance for others, 78% indicated that additional digital skills training would be beneficial [14].

We also found a psychosis-specific decrease in active social media posting. Social cognitive impairments and negative symptoms may contribute to this finding. This interpretation is supported by a study by Rehki et al. showing that an increase in severity of negative symptoms is associated with a lower likelihood of social media use [15]. In that study, they suggested that social interactions via social media should be considered in the clinical evaluation of individuals with schizophrenia, as it is a prominent form of communication and fostering relationships. While individuals with CR may also experience significant negative symptoms, our results suggest that these are not reflected in significant changes in social media posting and access rates. Although youths with PD post significantly less frequently than youths with CR and PF, individuals with PD nevertheless access social media at a similar rate, which provides a potential avenue for intervention. To our knowledge, no other study has distinguished between social media access and posting, so we are the first to report a decrease in social media posting for individuals with psychotic disorders while there was no difference in level of passive use of social media. This effect did not appear with mood disorders, ADHD, and anxiety, so therefore appears to be specific to psychosis.

Limitations of this study include sampling of one geographical area and limited sample size. Socioeconomic status may also influence access to technology but was not consistently measured in this sample. It is possible that patients with access to a specialized psychosis treatment center and those who volunteer for research participation may have higher access to technology than the general population of people with psychosis. Moreover, our survey only included two social media platforms, Facebook and Twitter. Future studies may consider including access and usage of other social media platforms that have recently become more popular among youths, as well as questions about their willingness to share their digital access and to receive therapeutic interactions via a digital platform. Objective usage statistics and other metadata may also provide useful signals. Privacy risks and concerns may also be technical as well as subjective barriers to implementation of such interventions. These were not addressed with the current study.

## Conclusions

Overall, our findings encourage further development of internet and social media-based interventions and monitoring for young people with psychosis and youths at clinical risk for psychosis. There appears to be no significant “technology gap” for young people with psychotic disorders relative to young people without psychosis. Lower active engagement may reflect impairments in social cognition and functioning. Notably, access to technology does not mean that digital health interventions will be ultimately efficacious or implementable – however, the availability of technology to young people with psychosis does provide some basis for feasibility. Future studies are needed to directly evaluate the efficacy and usability of digital health and social media-based strategies for intervention and assessment, while considering additional potentially harmful effects related to privacy concerns and increased time spent on social media.

## Data Availability

All data produced in the present study are available upon reasonable request to the authors.

## Acknowledgements

We thank the participants and their families for their collaboration. We also thank Lyndsay Schmidt, Zeeshan Huque, LaTonya McCurry, Victoria Pietruszka, Kosha Ruparel, Thomas Hohing, and Eehwa Ung from the University of Pennsylvania.

## Abbreviations

CSC: Coordinated Specialty
CR: Clinical risk
LiBI: Lifespan Brain Institute
PERC: Psychosis Evaluation and Recovery Center
PD: Psychosis disorder
PF: Psychosis-free
SIPS: Structured Interview for Prodromal Syndromes

